# Artificial Intelligence Significantly Improves Adenoma Detection Rate but Does Not Affect Polyp Detection Rate in Colonoscopy: A Propensity Score Matching Study

**DOI:** 10.1101/2025.11.09.25339868

**Authors:** Han Byul Lee, John Mayen Ruben, Byeong Cheol Jeong, Jun Sik Yoon, Seung Jung Yu, Eun Jeong Choi, Dong Woo Kim, Nguyen Quang Thu, David Lee, Soonwhan Kang, Jaeyoung Lee, Eunhye Kang, Nguyen Phuoc Long, Hong Sub Lee

## Abstract

**Background:** Colorectal cancer (CRC) remains a major cause of cancer-related morbidity and mortality worldwide. Endoscopy and adenoma removal are effective in reducing the incidence of CRC. Recent advances in artificial intelligence (AI)-assisted endoscopy have demonstrated the potential to improve detection outcomes. This study aimed to evaluate the effectiveness of AI-assisted endoscopy in improving adenoma detection rate (ADR) and polyp detection rate (PDR) during colonoscopy in a real-world clinical setting.

**Methods:** A total of 824 colonoscopies between August 2022 and February 2024 at Inje University Busan Paik Hospital were included in the study. Patients were divided into two groups: AI CAD-assisted colonoscopy (N = 393) and conventional colonoscopy (N = 431). Propensity score matching was then performed using a 1:1 nearest-neighbor algorithm, balancing key covariates, including age, sex, BMI, ASA score, bowel preparation quality, and the ratio of expert endoscopists. Ultimately, 786 patients (393 per group) were included in the final comparative analysis. Logistic regression analyses were used to evaluate the association between AI CAD-assisted colonoscopy and ADR and PDR, adjusting for potential confounders.

**Results:** ADR was significantly higher in the AI CAD-assisted group (41.5%) compared to the No-AI CAD group (34.4%) (adjusted OR = 1.380; 95% CI: 1.012–1.885; P = 0.042). PDR was higher in the AI CAD-assisted group (53.2% vs. 46.1%), but the difference was not statistically significant (OR = 1.312; 95% CI: 0.971–1.774; P = 0.077). Older age and higher BMI were positively associated with ADR, while male sex and higher ASA scores were negatively associated.

**Conclusions:** AI-assisted CAD colonoscopy was independently associated with improved ADR after adjustment for potential confounders. While the increase in PDR was not statistically significant, the findings support the clinical utility of AI CAD. Larger multicenter prospective studies are warranted to validate these findings and guide the integration of AI tools into routine endoscopic practice.

## INTRODUCTION

Colorectal cancer (CRC) ranks third in incidence and second in mortality among all cancers worldwide [1, 2]. In South Korea, colorectal cancer (CRC) is the third most common cancer [3], and the third leading cause of cancer-related deaths [4]. Because CRC often presents with symptoms such as occult blood, rectal bleeding, abdominal pain, and changes in bowel habits, mainly at advanced stages (Stage III or IV), early detection through screening methods such as blood tests [5], microsatellite instability testing [6], and colonoscopy [7], is essential. Detecting precancerous lesions such as adenomas and serrated lesions through colonoscopy and removing them with polypectomy is key to reducing mortality [8]. Among the identified risk factors, the typical precancerous lesions found in histopathological examinations of colorectal polyps are adenomas and serrated lesions. The adenoma-carcinoma pathway accounts for 70–90% of CRC cases, the serrated neoplasia pathway accounts for 10–20%, and microsatellite instability, such as in Lynch syndrome, contributes 2–7% [8]. Polyps found during screening colonoscopy undergo histopathological examination, and precancerous lesions are removed with polypectomy to prevent progression to colorectal cancer [9, 10].

A large-scale, multicenter randomized trial published in 2022 involving adult men and women aged 55 to 64 years from Poland, Norway, Sweden, and the Netherlands showed that the 10-year cumulative risk of colorectal cancer was 0.98 (95% CI, 0.86–1.09) in the group that underwent screening colonoscopy, compared to 1.20 (95% CI, 1.10–1.29) in the group that received only usual care, which corresponds to an 18% relative risk reduction (risk ratio, 0.82; 95% CI, 0.70–0.93) [11]. Furthermore, studies have demonstrated a strong link between higher detection rates of polyps and adenomas and a lower incidence of colorectal cancer after colonoscopy, highlighting the importance of identifying and removing adenomas during screening [12]. A growing body of evidence has consistently demonstrated that polypectomy performed during colonoscopy is effective in preventing the development of colorectal cancer.

In Korea, screening colonoscopy is recommended every five years for adults aged 50 and older [13]. The U.S. Preventive Services Task Force has recently updated its guidelines to recommend starting screening at age 45, lowering the age by five years compared to earlier recommendations [14]. Importantly, the adenoma detection rate (ADR) has long been established as a key quality indicator for colonoscopy [15], with guidelines recommending a minimum ADR of 35% (40% in men and 30% in women) when performed in asymptomatic individuals aged 45 years and older [16]. A multicenter study published in 2022 demonstrated a significant association between a median ADR of ≥ 28.3% and a reduced risk of colorectal cancer compared to lower ADRs [12]. Additionally, to ensure accurate detection and improve ADR, a minimum withdrawal time of six minutes is recommended.

The cognitive demands on gastroenterologists during real-time endoscopic procedures are immense, requiring them to process a vast amount of visual data, often exceeding 30 images per second, while making immediate decisions about diagnosis and treatment. This growing cognitive load has led to the development of deep learning-based artificial intelligence (AI) systems, particularly in computer vision, to assist in these complex tasks. AI computer-aided detection (CAD) is a gastrointestinal endoscopy AI software medical device that assists in the easy and accurate detection of lesions, regardless of their size and shape, through deep neural network-based learning. AI CAD displays detected polyps in real-time during endoscopic examinations, enabling healthcare professionals to make immediate medical judgments and take necessary actions. Since 2019, prospective randomized trials have demonstrated that CAD in colonoscopy significantly improves both the polyp detection rate (PDR) and ADR [17]. These systems typically function by highlighting suspicious polyps on the endoscopic monitor with “alert boxes,” thereby aiding the endoscopist’s visual inspection. The current limitations of these systems, however, often lie in their reduced sensitivity for smaller or more subtle polyps. Future research aims to enhance the detection of flat lesions and improve the real-time differentiation between benign and precancerous polyps. A major challenge in developing AI systems is the initial investment cost. Nonetheless, it will become more efficient compared to the time-consuming and labor-intensive process of traditional colonoscopy [18].

Since 2019, many international studies have consistently shown that AI-assisted colonoscopy improves detection rates. In a randomized controlled trial, Wang et al. reported significantly higher ADR and PDR with an AI-assisted system called EndoScreener [19]. These findings were further supported by a subsequent double-blinded RCT by the same group in 2020, which also demonstrated significant improvements in both ADR and PDR [20]. Likewise, a randomized trial examining the efficacy of real-time CAD for colorectal neoplasia found a significantly higher ADR with the GI-Genius system compared to conventional colonoscopy [21]. The benefit of AI has also been observed in reducing miss rates. A considerably lower adenoma miss rate in the AI-assisted group compared to the conventional group in the CADeT-CS Trial [22]. Similarly, a 2022 multicenter study in Japan utilizing the EndoBRAIN-EYE system showed significant increases in both ADR and PDR [23]. However, the impact of AI CAD on ADR and PDR in real-life clinical settings remains to be elucidated.

This study aimed to rigorously assess the impact of a novel AI-assisted polyp detection system, i.e., endoscopy as an AI CAD, on ADR in a real-world setting in a major medical center in South Korea. Using a retrospective review of electronic medical records from our institution, we compared ADR and PDR between colonoscopies performed with and without AI CAD assistance. This investigation provided robust evidence supporting the benefits of integrating AI into endoscopic practice to improve colorectal cancer screening outcomes.

## MATERIAL AND METHODS

### Population and Study Design

This study was conducted at the Endoscopy Center within the Department of Internal Medicine at Inje University Busan Paik Hospital. The study was approved by the Institutional Review Board of Inje University Busan Paik Hospital (BPIRB2023-11-11). All procedures and data analyses were performed in accordance with the ethical guidelines outlined in the Declaration of Helsinki.

Medical records for all colonoscopies performed between August 2022 and February 2024 were retrospectively reviewed. A total of 1,286 patients who underwent colonoscopy were initially enrolled. Patients were excluded if they were under 18 years of age, were pregnant, had poor bowel preparation (Boston Bowel Preparation Scale score < 6), or had a history of familial adenomatous polyposis (FAP), inflammatory bowel disease (IBD), or gastrointestinal cancer or surgery. Patients with incomplete baseline data were also excluded. Clinical outcomes were compared between two patient groups: those who underwent colonoscopy with AI CAD assistance and those who underwent conventional colonoscopy without AI assistance. The primary and secondary endpoints of the study were the ADR and PDR, respectively. These quality indicators were extracted from the medical records and were evaluated for differences between the two groups.

### Data Collection Procedure

Out of 1,286 colonoscopy records, 462 records were excluded, leaving 824 records available for analysis. To minimize selection bias, 1:1 nearest-neighbor propensity score matching (PSM) was performed using the MatchIt package to adjust for potential confounding variables [24]. The covariates included in the model were age, gender, BMI, ASA score, BBPS, and the ratio of expert endoscopists. After PSM, 786 colonoscopy records remained. AI CAD-assisted colonoscopies (N = 393) and non-AI CAD-assisted colonoscopies (N = 393) were identified following the matching (**Figure 1**). Of the 462 excluded cases, reasons included being under 18 years old, pregnancy, poor bowel preparation (Boston Bowel Preparation Scale < 6), a family history of familial adenomatous polyposis (FAP), a personal history of inflammatory bowel disease (IBD), and a history of gastrointestinal surgery.

**Figure 1.**
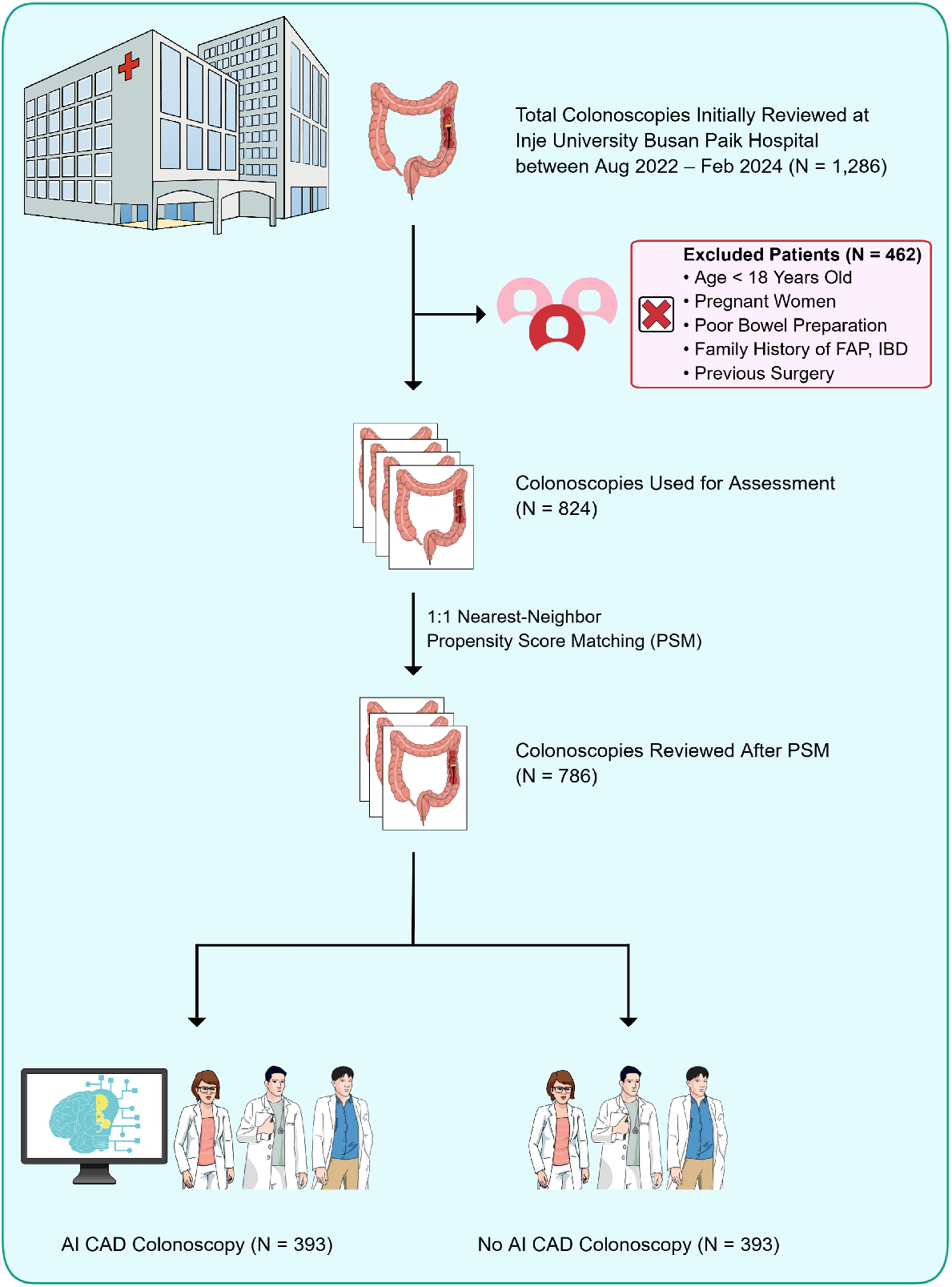
Flow chart for the selection of study participants. Abbreviations: AI CAD, Artificial intelligence computer-aided detection; FAP, Familial adenomatous polyposis; IBD, Inflammatory bowel disease; PSM: Propensity Score Matching.

The variables selected for baseline characteristics included age, sex, body mass index (BMI), obesity status, American Society of Anesthesiologists (ASA) physical status classification score, Boston Bowel Preparation Scale (BBPS), examiner proficiency in colonoscopy, and indications for colonoscopy. Obesity status was categorized as non-obese (BMI < 25) or obese (BMI ≥ 25). The ASA score was 1 for a healthy physical status and 2 for mild systemic disease. The BBPS scores were grouped into two categories: scores of 3–5 represented regular bowel preparation, and 6–9 represented good bowel preparation. Colonoscopy proficiency was defined as inexperienced (less than 1 year of colonoscopy practice) or experienced (1 year or more of colonoscopy practice). Indications for colonoscopy were classified into three categories: (1) surveillance after polypectomy, (2) symptomatic examination, and (3) screening without prior disease history.

### Statistical Analysis

Missing values were handled using listwise deletion. Continuous variables are reported as median with interquartile range (IQR), and categorical variables are presented as frequencies and percentages. To assess the normality of the distributions of continuous variables (e.g., age and BMI), the Shapiro-Wilk test was used. Because non-normal distributions were indicated (P < 0.05), the Mann-Whitney U test was used to compare these variables between the two groups. For categorical variables, including sex, obesity status, ASA score, Boston Bowel Preparation Scale (BBPS), indications for colonoscopy, and examiner proficiency, chi-square and Fisher’s exact tests were utilized to evaluate statistical differences in univariate analyses. A multivariable logistic regression analysis was performed with variables that demonstrated statistical significance in univariate analyses. The results were reported as odds ratios (ORs) with 95% confidence intervals (CIs) and corresponding P-values. All statistical analyses were performed using R software (version 4.2.1). All statistical tests were two-tailed, and P-values less than 0.05 were considered statistically significant.

## RESULTS

### Baseline Characteristics of the Study Before and After Propensity Score Matching

A total of 824 patients who underwent colonoscopy were included in the study. Before PSM, baseline characteristics were compared between the AI CAD-assisted colonoscopy group and the conventional colonoscopy group. There were no significant differences in age, gender, body mass index (BMI), obesity status, or BBPS between the groups. However, a significant imbalance was observed in the American Society of Anesthesiologists (ASA) score and the percentage of procedures performed by expert endoscopists (**Supplementary Table 1**). Following 1:1 PSM, a final cohort of 786 patients was established, with 393 patients in each group. After matching, all baseline characteristics, including age, gender, BMI, ASA score, BBPS, and expert endoscopist ratio, were well balanced between the groups (P > 0.05 for all comparisons, **Table 1**).

**Table 1.**
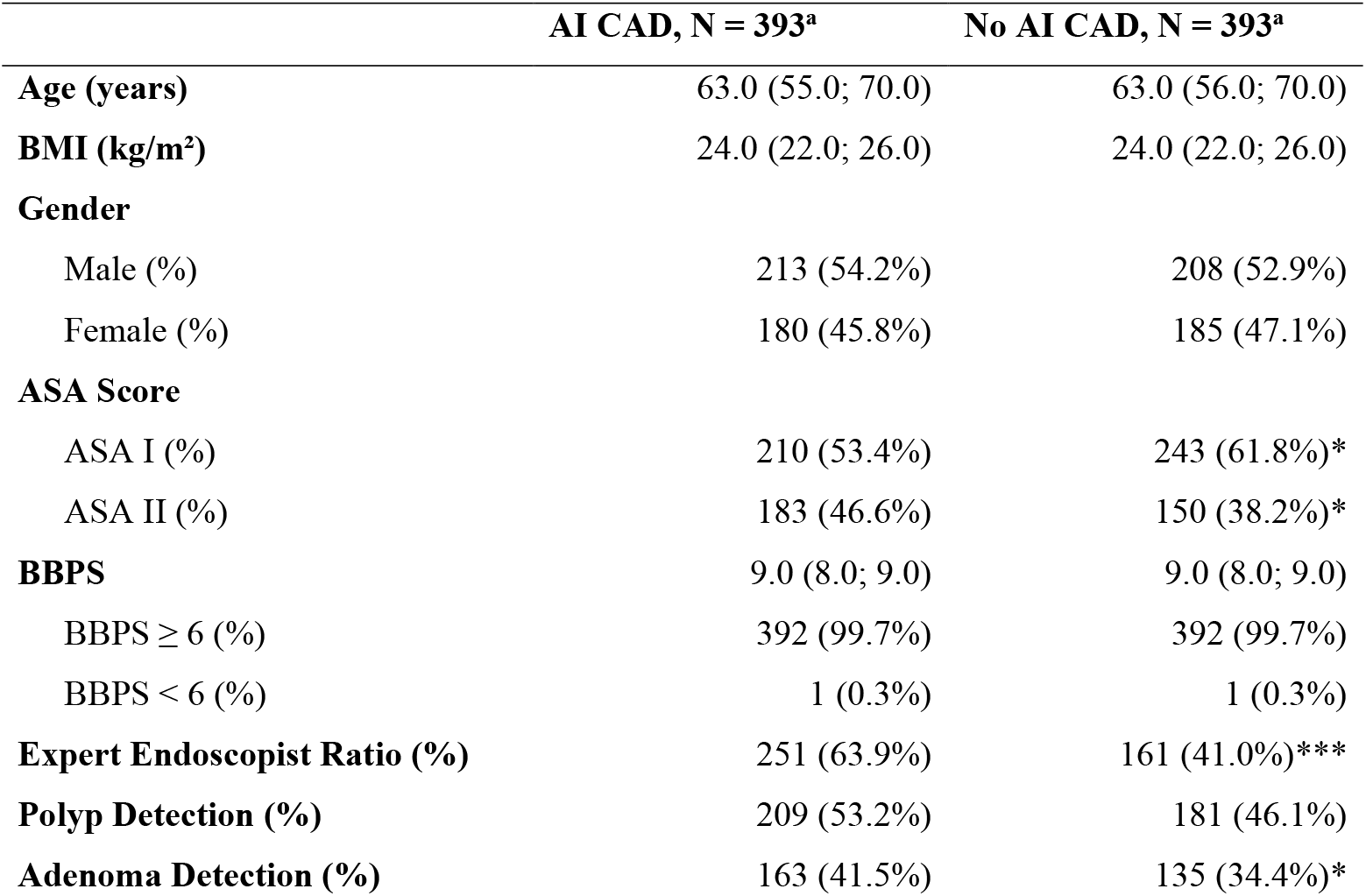
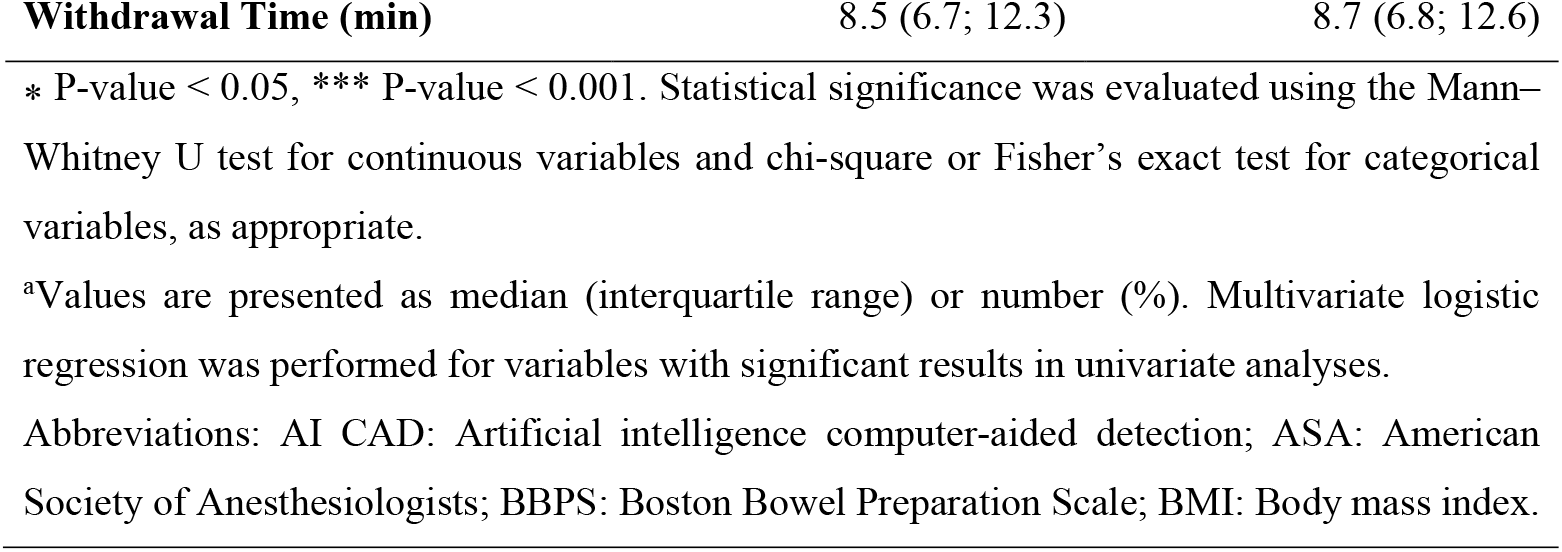
Baseline Characteristics After Propensity Score Matching (N = 786).

### Adenoma Detection Rate Was Significantly Higher in the AI CAD Group

We first investigated the ADR of endoscopy with and without the assistance of AI CAD. Before PSM, the ADR was significantly higher in the AI CAD group compared to the conventional colonoscopy (No-AI CAD) group (41.5% vs. 33.6%; N = 824, P=0.024). The multivariable logistic regression analysis indicated that the use of AI CAD was significantly associated with an increased ADR (adjusted OR, 1.436; 95% CI, 1.056–1.956; P = 0.021) (**Supplementary Table 2**). Other factors positively associated with ADR included BMI and age. On the contrary, being male and having a higher ASA score were negatively associated with ADR.

**Table 2.**
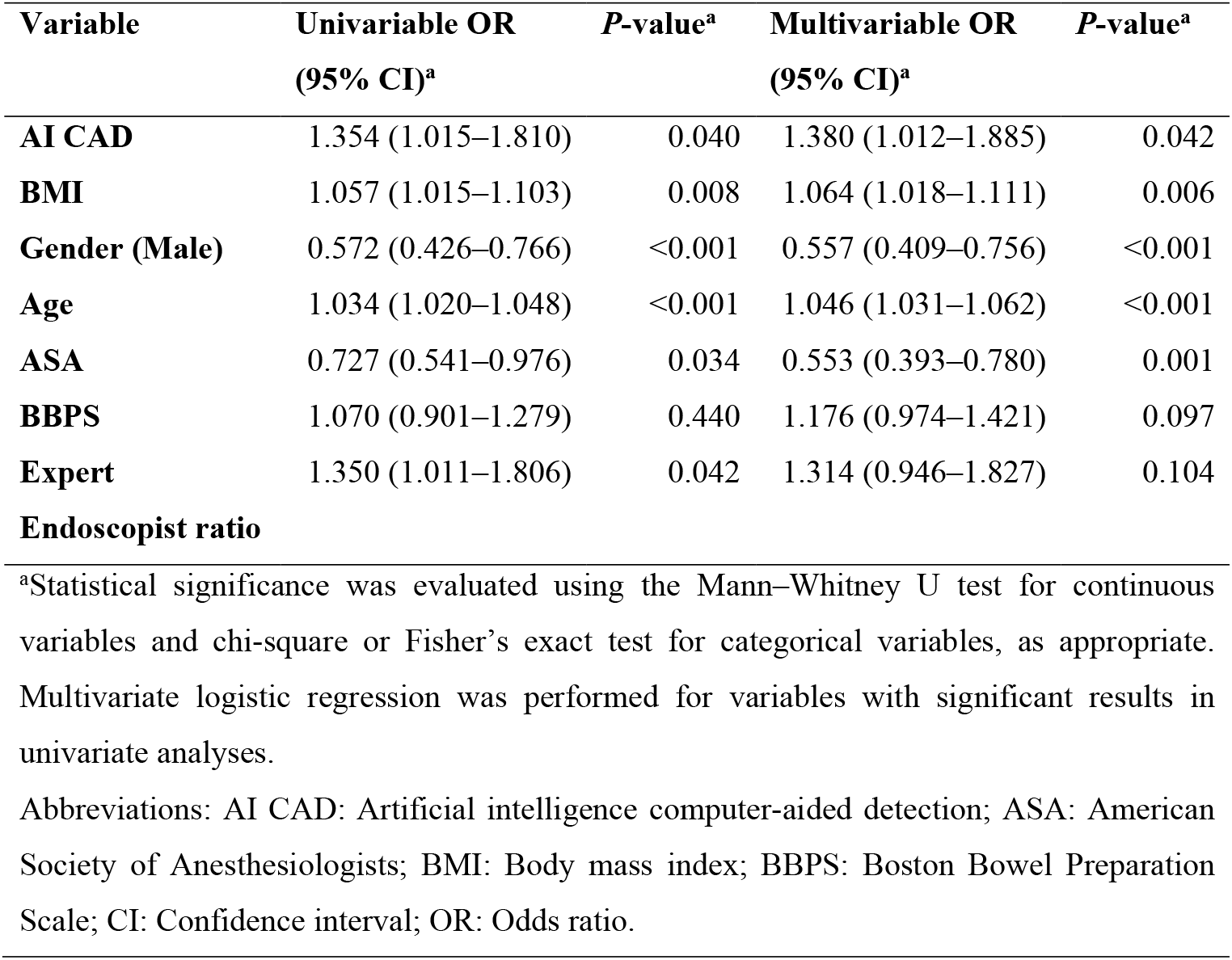
Logistic Regression Analysis Evaluating the Effect of AI CAD on Adenoma Detection Rate (ADR) in 786 Patients. Abbreviation: AI CAD: Artificial intelligence computer-aided detection.

After 1:1 propensity score matching, the ADR in the AI CAD group remained significantly higher (41.5%) than in the No-AI CAD group (34.4%) (N = 786, P = 0.047). The multivariable logistic regression analysis further confirmed that AI CAD use was independently associated with a higher ADR (adjusted OR, 1.380; 95% CI, 1.012–1.885; P = 0.042) (**Table 2**). Similar to the pre-PSM analysis, BMI and age were positively associated with ADR, and male gender and a higher ASA score were negatively associated.

### Polyp Detection Rate Was Not Significantly Higher in the AI CAD Group

Following the ADR, we further examined if AI CAD significantly improve the PDR in endoscopy before and after PSM. Before PSM, logistic regression analysis demonstrated a significant association between AI CAD use and a higher PDR (OR, 1.354; 95% CI, 1.006– 1.826; P = 0.046) (**Supplementary Table 3**). Factors positively associated with PDR were age and BMI, whereas being male and a higher ASA score were negatively associated. After PSM, the PDR was higher in the AI CAD group (53.2%) compared with the No-AI CAD group (46.1%), though the difference was not statistically significant (OR, 1.312; 95% CI, 0.971– 1.774; P = 0.077) (**Table 3**). Consistent with the pre-PSM findings, BMI and age were positively associated with PDR, while male gender and a higher ASA score were negatively associated.

**Table 3.** Logistic Regression Analysis Evaluating the Effect of AI CAD on Polyp Detection Rate (PDR) After Propensity Score Matching (N = 786). Abbreviation: AI CAD: Artificial intelligence computer-aided detection.

| Variable | OR (95% CI) | P-value<sup>a</sup> |
| --- | --- | --- |
| AI CAD | 1.312 (0.971–1.774) | 0.077 |
| BMI | 1.069 (1.025–1.114) | 0.002 |
| Gender (Male) | 0.522 (0.387–0.703) | <0.001 |
| Age | 1.029 (1.015–1.043) | <0.001 |
| ASA | 0.517 (0.377–0.706) | <0.001 |
| BBPS | 1.105 (0.924–1.326) | 0.276 |
| Expert Endoscopist Ratio | 1.420 (1.034–1.954) | 0.031 |

<sup>a</sup>Statistical significance was evaluated using the Mann–Whitney U test for continuous variables and chi-square or Fisher’s exact test for categorical variables, as appropriate. Multivariate logistic regression was performed for variables with significant results in univariate analyses.
Abbreviations: AI CAD: Artificial intelligence computer-aided detection; ASA: American Society of Anesthesiologists; BMI: Body mass index; BBPS: Boston Bowel Preparation Scale; CI: Confidence interval; OR: Odds ratio.

## DISCUSSION

In this study, we evaluated the clinical utility of AI CAD regarding the ADR and PDR in colonoscopy. After applying propensity score matching to mitigate selection bias, we found a statistically significant increase in ADR with AI-assisted colonoscopy compared with conventional colonoscopy. Although the PDF was also higher in the AI group, this difference was not statistically significant. This improvement in ADR is particularly noteworthy. This finding aligns with prior studies, which have shown a similar increase of approximately 10% to 15% with AI-assisted colonoscopy [21, 25, 26]. The clinical significance of this finding is profound because an elevated ADR is a crucial quality metric directly linked to a reduced risk of interval colorectal cancer. Our results also suggest that AI-based systems can standardize detection quality, potentially reducing operator variability and helping endoscopists avoid missed lesions.

AI-based systems, particularly those utilizing deep learning, are being increasingly applied in colonoscopies to provide real-time assistance by detecting and highlighting polyps. By identifying lesions that might otherwise be missed, these systems have been shown in multiple studies to improve both PDR and ADR [20-23]. There may be significant differences in detection rates between experienced and inexperienced colonoscopists. This emphasizes the need to further investigate the influence of colonoscopy experience. In the present study, the expert endoscopist ratio, that is, the proportion of experienced colonoscopists (those with more than one year of experience), was lower in the conventional colonoscopy group compared to experienced colonoscopists in the AI-assisted group (37.4% vs 63.9% before PSM, 41.0% vs 63.9% after PSM). Subsequent large-scale studies should examine whether a higher proportion of experienced colonoscopists might yield a more pronounced increase in ADR and PDR when assisted by AI. It is important to note that one study with a high endoscopy rate (78%) performed by experienced endoscopists found that ADR was not significantly higher in the AI-assisted group [27]. Additionally, the adenoma miss rate was considerably lower among AI-assisted novice colonoscopists compared to those without AI support, demonstrating a clear advantage of AI-assisted endoscopy. The study also showed that AI-assisted novice colonoscopists were not inferior compared to the experts [28]. Our work using the PSM study design provides a fairer comparison of the clinical utility of AI CAD, indicating a significantly higher ADR but not PDR in the AI-assisted group. Our findings support the clinical use of AI to improve colonoscopy quality metrics and the potential benefits on the prognosis of the patients.

This study has several limitations that must be acknowledged. First, it was a retrospective, single-center study, which may limit the generalizability of our findings to other clinical settings. Second, although PSM was used to adjust for confounding variables, residual confounding from unmeasured factors, such as detailed withdrawal techniques or lesion morphology, could not be completely ruled out. Third, despite the relatively large overall sample size, subgroup analyses were limited by the number of procedures performed by each operator. We could not assess inter-examiner agreement and could only categorize colonoscopists by experience level rather than individual skill.

## CONCLUSIONS

This study found that AI-assisted colonoscopy with AI CAD was associated with a higher detection rate of adenomas, particularly after excluding symptomatic patients and applying propensity score matching. However, the increase in PDR was not statistically significant in the AI CAD-assisted group compared to the no AI-CAD-assisted group. These findings underscore the potential of deep learning-based AI to enhance colonoscopy outcomes, which could improve colorectal cancer prevention. Future research, including multi-center, prospective studies with larger sample sizes and a more comprehensive set of clinical variables, is warranted to validate these results and evaluate the broader applicability of AI CAD in routine clinical practice.

## Data Availability

All data produced in the present study are available upon reasonable request to the authors

## Acknowledgement

None.

