## Supplementary File for "Artificial Intelligence Significantly Improves Adenoma Detection Rate but Does Not Affect Polyp Detection Rate in Colonoscopy: A Propensity Score Matching Study"

[3]3F Onnuri Medical Internal Medicine, Busan 47392, Republic of Korea

[4]Graduate Institute of Biomedical Sciences, College of Medicine, Chang Gung University, Taoyuan 333, Taiwan

[5]Ainex Corporation, Seoul 0617, Republic of Korea

[6]Molecular Medicine Research Center, College of Medicine, Chang Gung University, Taoyuan 333, Taiwan

[*]These authors contributed equally to this work.

**SUPPLEMENTARY TABLES**

**Supplementary Table 1. Baseline Characteristics Prior to Propensity Score Matching (N = 824), Primary Outcome: ADR.** Abbreviations: ADR: Adenoma detection rate.

| | **AI CAD, N = 393[a]** | **No AI CAD, N = 431[a]** |
|---|---|---|
| **Age (years)** | 63.0 (55.5; 70.0) | 64.0 (56.5; 70.0) |
| **Gender** | | |
| Male (%) | 213 (54.2%) | 231 (53.6%) |
| Female (%) | 180 (45.8%) | 200 (46.4%) |
| **BMI (kg/m²)** | 23.0 (22.0; 26.0) | 24.0 (22.0; 26.0) |
| **ASA Score** | | |
| ASA I (%) | 210 (53.4%) | 281 (65.2%)** |
| ASA II (%) | 183 (46.6%) | 150 (34.8%)** |
| ASA III (%) | 0 (0%) | 0 (0%) |
| **BBPS** | | |
| BBPS ≥ 6 (%) | 392 (99.7%) | 429 (99.5%) |
| BBPS < 6 (%) | 1 (0.3%) | 2 (0.5%) |
| **Expert Endoscopist ratio (%)** | 251 (63.9%) | 161 (37.4%)*** |
| **Withdrawal Time (min)** | 8.5 (6.7; 12.3) | 8.6 (6.7; 12.3) |
| **Polyp Detection Rate (%)** | 209 (53.2%) | 195 (45.2%)* |
| **Adenoma Detection Rate (%)** | 163 (41.5%) | 145 (33.6%)* |

* P-value < 0.05, ** P-value < 0.01, *** P-value < 0.001. Statistical significance was evaluated using the Mann–Whitney U test for continuous variables and chi-square or Fisher's exact test for categorical variables, as appropriate.

[a]Values are presented as median (interquartile range) or number (%). Multivariate logistic regression was performed for variables with significant results in univariate analyses.

Abbreviations: AI CAD: Artificial intelligence computer-aided detection; ASA: American Society of Anesthesiologists; BBPS: Boston Bowel Preparation Scale; BMI: Body mass index.

**Supplementary Table 2. Logistic Regression Analysis Evaluating the Effect of AI CAD on Adenoma Detection Rate (ADR) in 824 patients.** Abbreviation: AI CAD: Artificial intelligence computer-aided detection.

| Variable | Univariable OR (95% CI) | *P*-value[a] | Multivariable OR (95% CI) | *P*-value[a] |
|---|---|---|---|---|
| **AI CAD** | 1.398 (1.053–1.857) | 0.020 | 1.436 (1.056–1.956) | 0.021 |
| **BMI** | 1.060 (1.018–1.105) | 0.005 | 1.066 (1.021–1.113) | 0.003 |
| **Gender** | 0.589 (0.441–0.785) | < 0.001 | 0.575 (0.425–0.779) | < 0.001 |
| **Age** | 1.033 (1.020–1.047) | < 0.001 | 1.046 (1.031–1.062) | < 0.001 |
| **ASA** | 0.763 (0.570–1.019) | 0.068 | 0.579 (0.419–0.798) | 0.001 |
| **BBPS** | 1.101 (0.933–1.307) | 0.262 | – | – |
| **Expert Endoscopist ratio** | 1.395 (1.051–1.853) | 0.021 | 1.385 (1.002–1.918) | 0.049 |

[a]Statistical significance was evaluated using the Mann–Whitney U test for continuous variables and chi-square or Fisher's exact test for categorical variables, as appropriate. Multivariate logistic regression was performed for variables with significant results in univariate analyses.

Abbreviations: AI CAD: Artificial intelligence computer-aided detection; ASA: American Society of Anesthesiologists; BMI: Body mass index; BBPS: Boston Bowel Preparation Scale; CI: Confidence interval; OR: Odds ratio.

**Supplementary Table 3. Logistic Regression Analysis Evaluating the Effect of AI CAD on Polyp Detection Rate (PDR) in 824 Patients.** Abbreviation: AI CAD: Artificial intelligence computer-aided detection.

| **Variable** | **OR (95% CI)** | ***P*-value[a]** |
|---|---|---|
| **AI CAD** | 1.354 (1.006–1.826) | 0.046 |
| **BMI** | 1.071 (1.027–1.117) | 0.001 |
| **Gender** | 0.530 (0.395–0.708) | < 0.001 |
| **Age** | 1.029 (1.015–1.043) | < 0.001 |
| **ASA** | 0.541 (0.396–0.735) | < 0.001 |
| **BBPS** | 1.120 (0.943–1.334) | 0.201 |
| **Expert Endoscopist Ratio** | 1.497 (1.095–2.051) | 0.012 |

[a]Statistical significance was evaluated using the Mann–Whitney U test for continuous variables and chi-square or Fisher's exact test for categorical variables, as appropriate. Multivariate logistic regression was performed for variables with significant results in univariate analyses.

Abbreviations: AI CAD: Artificial intelligence computer-aided detection; ASA: American Society of Anesthesiologists; BMI: Body mass index; BBPS: Boston Bowel Preparation Scale; CI: Confidence interval; OR: Odds ratio.
